# Sociodemographic inequalities in COVID-19 booster vaccination in Peru: a GINI index decomposition analysis

**DOI:** 10.1101/2023.06.10.23291225

**Authors:** Claudio Intimayta-Escalante, Gustavo Tapia-Sequeiros, Daniel Rojas-Bolivar

## Abstract

**Background:** COVID-19 vaccine coverage, especially in booster doses, remains a significant issue in Peru. This study aimed to analyze the social determinants that affect vaccine coverage and evaluate sociodemographic inequalities in COVID-19 booster dose vaccination in Peru.

**Methods:** An ecological study was conducted with 196 provinces in Peru. GINI index decomposition analysis was performed to assess the inequality of immunization coverage in these provinces, considering sociodemographic characteristics as sources of inequality (sex, age group, educational level, residence area, and ethnic group). The Oaxaca-Blinder method was utilized to decompose the GINI index into Sk (composition effect), Gk (redistribution effect), Rk (differential effect), share value, and percentage change. Bootstrap method based on percentiles was employed to determine 95% confidence interval values for each GINI index and percentage change in the decomposition analysis.

**Results:** A total of 196 Peruvian provinces were evaluated. Among these, 59.74% had higher education, while 10.37% had no education or only elementary education. White or mestizo individuals accounted for 51.62% of the population. The GINI index decomposition analysis, six months after the start of the third and fourth dose vaccination campaign, revealed higher Sk values for the white or mestizo ethnic group (Sk= 6.46 vs Sk= 3.03), people living in rural (Sk= 6.63 vs Sk= 2.76) or urban areas (Sk= 5.88 vs Sk= 2.76), and those aged between 30 and 64 years (Sk= 6.84 vs Sk= 3.20). The highest Gk values were observed for the Aymara (Gk= 0.92 vs Gk= 0.92), Afro-Peruvian (Gk= 0.61 vs Gk= 0.61), and Quechua (Gk= 0.53 vs Gk= 0.53) ethnic groups, in contrast to the white or mestizo group (Gk= 0.34 vs Gk= 0.34). Furthermore, Rk exhibited high positive values for individuals with university or postgraduate education (Rk= 0.59 vs Rk= 0.44) and those living in urban areas (Rk= 0.34 vs Rk= 0.28). Conversely, individuals living in rural areas (Rk= -0.34 vs Rk= -0.28), those with elementary education (Rk= -0.46 vs. Rk= -0.36), and those with no education or only preschool education (Rk= -0.41 vs. Rk= -0.32) displayed high negative Rk values.

**Conclussion:** Sociodemographic inequalities exist in the coverage of third and fourth booster doses against COVID-19 in Peruvian provinces, particularly concerning geographic location, ethnicity, and educational level.

## BACKGROUND

The COVID-19 pandemic has impacted all the world, the number of people infected, and dead has dramatically increased (1), especially in low-and middle income-countries, where the number of deaths is above of 6.9 million (2). Several factors have influenced on the COVID-19 transmissibility and mortality rate, such as health system performance, social inequalities, and access to vaccines (3-5).

Also, it has been reported that vaccinations significantly reduced the mortality rate as well as the virus transmission (6,7). However, inequal access affects the vaccines distribution and therefore the incidence and mortality rates could be higher in more vulnerable populations, such as ethnic groups, people living in poverty or living in rural areas (8–10). In some countries, like the United States, inequities in access to vaccinations was higher among non-white ethnic groups (black or Hispanic people) than white people (10,11), while in Canada, the proportion of people vaccinated was higher in urban areas in comparison con rural areas (8,9).

Absolute differences in vaccination coverage rate between groups has been used as an indicator in health inequality studies (12,13). However, the Gini coefficient has been described as the most used method to estimate the unequal distribution of a resource or service for specific populations (11,14). Nevertheless, it only has been used in some studies about health inequalities in low -and middle-income-countries (15,16).

COVID-19 vaccine coverage remains as a significant problem in Peru, particularly in booster doses campaign, and its consequences are still can increase the number of deceases as well as the people hospitalized in intensive care units (17).Therefore, to analyze the social determinants which affects the vaccine coverage, the aim of this study was evaluate sociodemographic inequalities in vaccination with COVID-19 booster doses in Peru using GINI index decomposition analysis.

## METHODS

### Study Design

An ecological study was conducted. Data of 196 provinces of Peru were analyzed. The objective was to collect information of the number of inhabitants over 12 years old who received the COVID-19 third and fourth booster doses in each province from the beginning of the vaccination campaign to 12 months later. Peruvian provinces data were obtained from the website of the Ministry of Health (https://www.minsa.gob.pe/reunis/data/vacunas-covid19.asp).

### Definition of Variables

The sociodemographic characteristics of Peruvian residents aged 15 and above were sex, age distribution (15-19, 20-29, 30-64, and over 65 years), education level (none education or only preschool education, elementary, high school, and university or post-graduate), residence area (urban or rural), and ethnicity (mestizo or white, quechua, aimara, afro-peruvian, and other groups). The data source was the 2017 National Census in Peru, which was carried out by the National Institute of Statistics and Informatics (https://censo2017.inei.gob.pe/resultadosdefinitivos-de-los-censos-nacionales-2017/).

COVID-19 positive rate and vaccination coverage data at six and twelve months after the beginning of the booster vaccination campaign was collected from the Open Data National Platform in Peru (https://www.datosabiertos.gob.pe/dataset/casos-positivos-por-covid-19-ministerio-desalud-minsa).

### Statistical Analysis

Statistical analysis was performed using STATA software version 17.0. Descriptive analysis included mean percentage of each sociodemographic characteristic and their respective 95% confidence intervals.

Analysis of inequality of COVID-19 booster dose vaccination coverage used cumulative percentage of population in each province. Lorenz’s curves were used to visualize the distribution of inequality and to estimate the GINI index, which reflects the equitable distribution of a resource or service, where a value of 0 indicates perfect equity and a value of 1 indicates absolute inequality (18).

However, since the GINI index is an indicator of absolute inequality, we used a decomposition approach with the Oaxaca-Blinder method. This method addresses various characteristics as sources of inequality in a population (19). The decomposition of the GINI index was carried out with the following formula: :

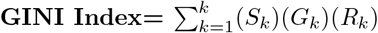

According to the formula, Sk (composition effect) is the contribution of each characteristic to total inequality. While Gk (redistribution effect) is the GINI index for each characteristic. As well, Rk (differential effect) is the correlation between inequality of booster doses coverage with the percentage values of each sociodemographic characteristic (20).

In addition, the Oaxaca-Blinder decomposition method estimates the share, which expresses the proportion of vaccination coverage for each characteristic included. The share value is used to estimate the compositional effect of each characteristic on overall GINI index value; as well as to determine the effect it could have given some scenarios (values above 0 reduce inequality, while values below 0 increase inequality). We also estimate the percentage change or %change, which expresses the change in GINI index due to a 10% of change in each characteristic evaluated.

Likewise, the Bootstrap method based on percentiles was used to generate simulate random samples and determine 95% confidence interval values for each GINI index and percentage change values in decomposition analysis.

### Ethical Aspects

This study used open available data from the Ministry of Health of Peru. Therefore, there was no intervention on humans. So, ethical board approval was not mandatory.

## RESULTS

In the 196 Peruvian provinces were evaluated. The mean percentage of men and women in provinces was similar (46.50% and 46.70%, respectively). Most inhabitants completed higher education (59.74%), while population without education or only elementary education was 10.37%. Half of the inhabitants in each province was between 30 and 64 (50.50%), and most provinces were in urban areas (53.51%). Most of the population identified themselves as white or mestizo (51.62%), followed by the Quechua group (34.79%), Aymara (4.01%), Afro-Peruvian (2.86%), and others (0.44%). The mean positivity rate of COVID-

19 in provinces has been very low from the beginning of vaccination campaign, it slightly increased from 13.94% at six months after the beginning of vaccination campaign to 17.33% at six months. However, the mean COVID-19 positivity rate raised when the fourth dose vaccination campaign started in provinces, reaching a percentage of 18.78% and 19.33% at six and twelve months, respectively (**Table 1**). The mean immunization coverage increased after the third dose from 13.38% at six months to 57.0% at twelve months. As well, mean immunization coverage increased from 21.20% at six months to 22.19% at twelve months (**Table 1**).

**Table 1.**
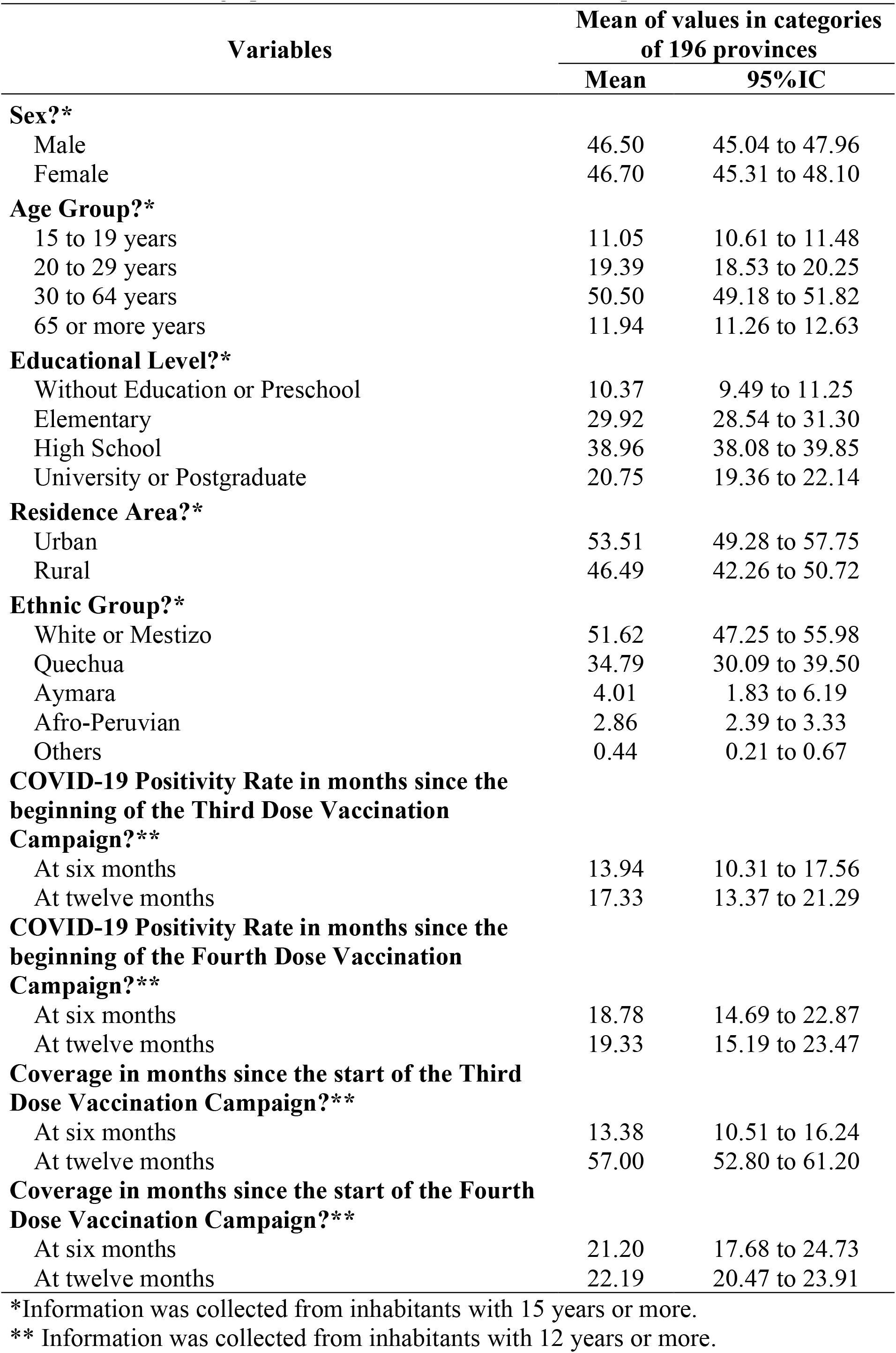
Sociodemographic characteristics of 196 Peruvian provinces

The overall GINI coefficient was 0.84 (95% CI: 0.80 to 0.88) during the first month of COVID-19 third dose immunization program. At six months after the beginning of vaccination campaign, vaccination coverage increased to 7.99% (95% CI: 7.34 to 8.65), and the GINI index decreased to 0.33 (95%IC: 0.30 to 0.35) (**Figure 1A**). Regarding COVID-19 fourth dose campaign, the Gini index was 0.47 (95%CI: 0.43 to 0.51) at the first month. Then, it declined to 0.33 (95%CI: 0.31 to 0.35) at six months, and 0.31 (95%CI: 0.29 to 0.32) at twelve months. On the other hand, vaccination coverage increased from 1% to 17% from the beginning to six months, while at twelve months, vaccination coverage rate was 22.19 (95% CI: 20.47 to 23.91) (**Figure 1B**).

**Figure 1.**
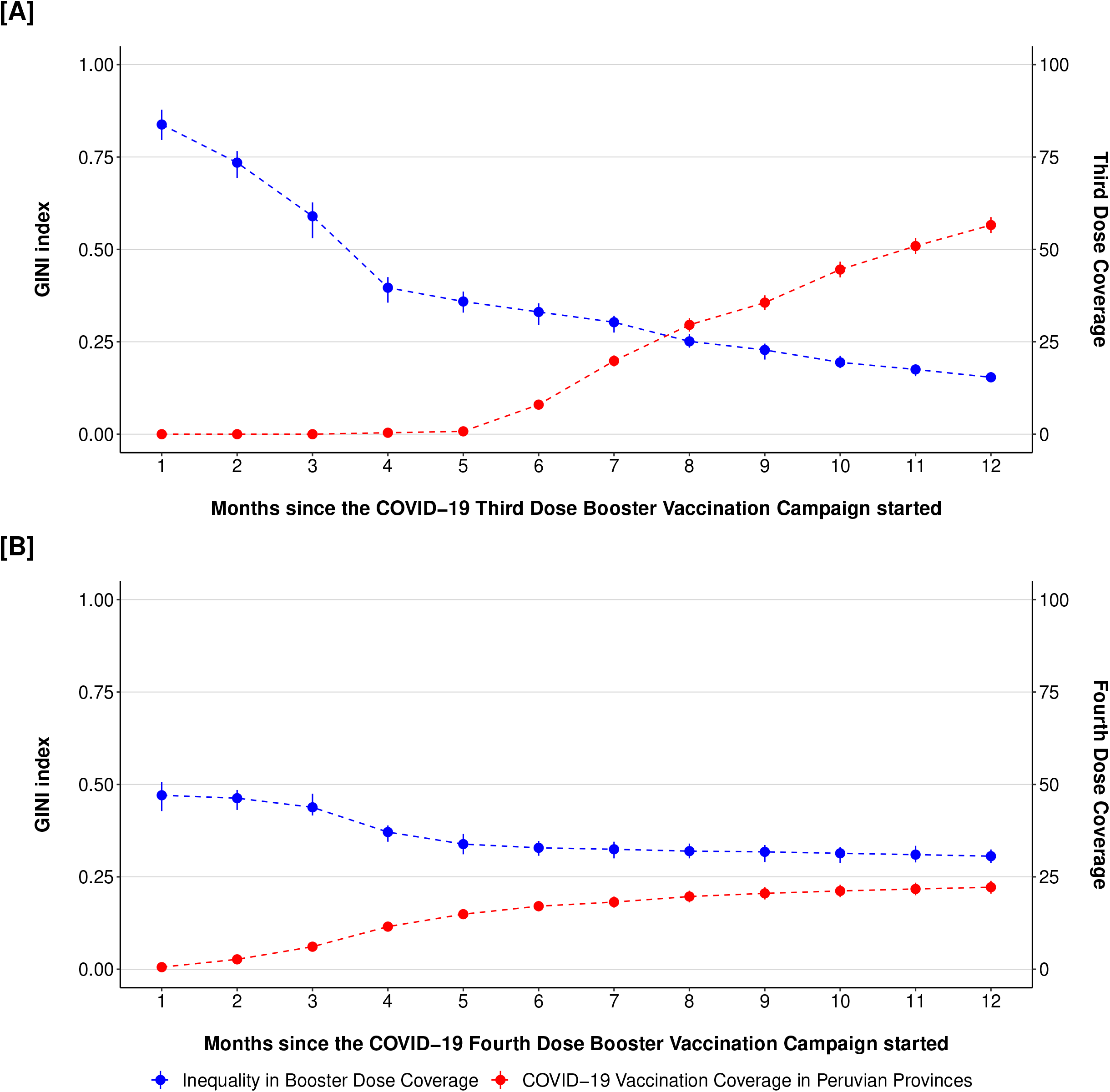
Evolution of the GINI coefficient and booster dose coverage for COVID–19 in the 196 Peruvian provinces from the beginning of the vaccination campaign until 12 months of age

Regarding Gini coefficient decomposition analysis at six months after the start of third dose vaccination campaign, the contribution of each sociodemographic characteristic to total inequality (Sk) was higher in white or mestizo ethnic group (Sk= 6.46), people living in rural (Sk= 6.63) or urban areas (Sk= 5.88), as well as the age group between 30 and 64 years old (Sk= 6.84), and for men and women (Sk= 6.23 and Sk= 6.28, respectively). In addition, it was identified that the GINI coefficient for each sociodemographic characteristic (Gk) was higher for the Aymara (Gk= 0.92), Afro-Peruvian (Gk= 0.61), and Quechua (Gk=0.53) ethnic groups, as opposed to the white or mestizo group (Gk= 0.34). Likewise, the correlation between the GINI index and the values of sociodemographic characteristics (Rk) showed high positive values for inhabitants with university or postgraduate education (Rk= 0.59), for people living in urban areas (Rk= 0.34) and for women in provinces (Rk= 0.10). Moreover, high negative values were identified for those living in rural areas (Rk= -0.34), population with elementary education (Rk= -0.46), and those with no education or only preschool education (Rk= -0.41). These trends remained consistent in the GINI index decomposition analysis at twelve months after the start of third dose vaccination campaign, however the values were low. Furthermore, another important factor in the inequality in vaccination coverage was the percentage of positivity in provinces; both six (Sk= 1.74, Gk= 0.71, and Rk= -0.05) and twelve (Sk= 0.31, Gk= 0.67, and Rk= -0.04) months after the beginning of the third dose vaccination campaign (**Table 2**).

**Table 2.**
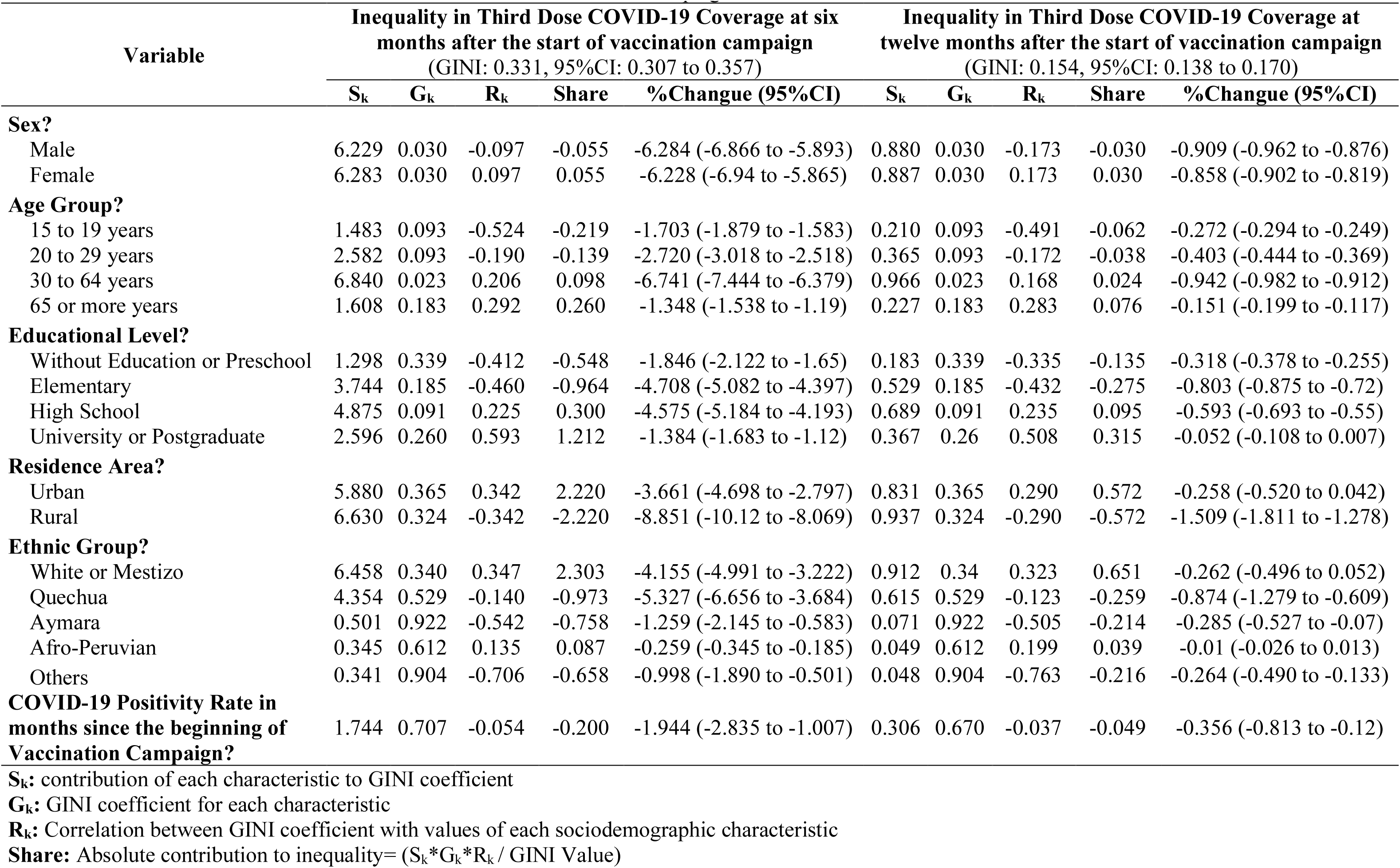
Decomposition of the GINI coefficient on inequality in coverage for the third booster dose for COVID-19 between sociodemographic characteristics at six and twelve months after the start of the vaccination campaign

**Table 3.** Decomposition of the GINI coefficient on inequality in coverage for the fourth booster dose for COVID-19 between sociodemographic characteristics at six and twelve months after the start of the vaccination campaign

| Variable | Inequality in Fourth Dose COVID-19 Coverage at six months after the start of vaccination campaign<br>(GINI: 0.329, 95%CI: 0.298 to 0.354) |  |  |  |  | Inequality in Fourth Dose COVID-19 Coverage at twelve months after the start of vaccination campaign<br>(GINI: 0.306, 95%CI: 0.282 to 0.325) |  |  |  |  |
| --- | --- | --- | --- | --- | --- | --- | --- | --- | --- | --- |
|  | S<sub>k</sub> | G<sub>k</sub> | R<sub>k</sub> | Share | %Change (95%CI) | S<sub>k</sub> | G<sub>k</sub> | R<sub>k</sub> | Share | %Change (95%CI) |
| Sex? |  |  |  |  |  |  |  |  |  |  |
| Male | 2.918 | 0.030 | -0.165 | -0.044 | -2.963 (-3.223 to -2.756) | 2.244 | 0.030 | -0.178 | -0.039 | -2.283 (-2.402 to -2.112) |
| Female | 2.944 | 0.029 | 0.165 | 0.044 | -2.899 (-3.185 to -2.686) | 2.263 | 0.030 | 0.178 | 0.039 | -2.224 (-2.368 to -2.049) |
| Age Group? |  |  |  |  |  |  |  |  |  |  |
| 15 to 19 years | 0.695 | 0.093 | -0.415 | -0.082 | -0.777 (-0.859 to -0.697) | 0.534 | 0.093 | -0.373 | -0.061 | -0.595 (-0.641 to -0.538) |
| 20 to 29 years | 1.210 | 0.093 | -0.144 | -0.05 | -1.259 (-1.390 to -1.148) | 0.930 | 0.093 | -0.151 | -0.043 | -0.973 (-1.024 to -0.88) |
| 30 to 64 years | 3.204 | 0.023 | 0.136 | 0.031 | -3.174 (-3.448 to -2.958) | 2.464 | 0.023 | 0.096 | 0.018 | -2.446 (-2.59 to -2.269) |
| 65 or more years | 0.753 | 0.183 | 0.241 | 0.101 | -0.652 (-0.732 to -0.589) | 0.579 | 0.183 | 0.248 | 0.086 | -0.493 (-0.569 to -0.433) |
| Educational Level? |  |  |  |  |  |  |  |  |  |  |
| Without Education or Preschool | 0.608 | 0.339 | -0.323 | -0.203 | -0.811 (-0.941 to -0.694) | 0.468 | 0.339 | -0.278 | -0.144 | -0.611 (-0.732 to -0.495) |
| Elementary | 1.754 | 0.185 | -0.364 | -0.36 | -2.114 (-2.362 to -1.95) | 1.349 | 0.185 | -0.334 | -0.273 | -1.621 (-1.739 to -1.467) |
| High School | 2.284 | 0.091 | 0.228 | 0.144 | -2.140 (-2.393 to -1.942) | 1.756 | 0.091 | 0.190 | 0.099 | -1.657 (-1.77 to -1.442) |
| University or Postgraduate | 1.216 | 0.260 | 0.435 | 0.419 | -0.797 (-0.903 to -0.681) | 0.935 | 0.260 | 0.399 | 0.318 | -0.618 (-0.679 to -0.5) |
| Residence Area? |  |  |  |  |  |  |  |  |  |  |
| Urban | 2.755 | 0.365 | 0.282 | 0.865 | -1.890 (-2.245 to -1.478) | 2.118 | 0.365 | 0.240 | 0.607 | -1.511 (-1.792 to -1.258) |
| Rural | 3.107 | 0.324 | -0.282 | -0.865 | -3.972 (-4.596 to -3.616) | 2.389 | 0.324 | -0.240 | -0.607 | -2.996 (-3.363 to -2.552) |
| Ethnic Group? |  |  |  |  |  |  |  |  |  |  |
| White or Mestizo | 3.026 | 0.340 | 0.361 | 1.130 | -1.896 (-2.596 to -1.302) | 2.327 | 0.340 | 0.340 | 0.879 | -1.448 (-1.791 to -1.126) |
| Quechua | 2.040 | 0.529 | -0.158 | -0.518 | -2.557 (-3.151 to -1.792) | 1.568 | 0.529 | -0.136 | -0.368 | -1.937 (-2.39 to -1.445) |
| Aymara | 0.235 | 0.922 | -0.648 | -0.428 | -0.663 (-1.053 to -0.309) | 0.181 | 0.922 | -0.662 | -0.361 | -0.541 (-0.869 to -0.273) |
| Afro-Peruvian | 0.162 | 0.612 | 0.185 | 0.056 | -0.106 (-0.150 to -0.072) | 0.124 | 0.612 | 0.168 | 0.042 | -0.083 (-0.105 to -0.057) |
| Others | 0.160 | 0.904 | -0.568 | -0.250 | -0.409 (-0.759 to -0.232) | 0.123 | 0.904 | -0.535 | -0.194 | -0.317 (-0.507 to -0.124) |
| COVID-19 Positivity Rate in months since the beginning of Vaccination Campaign? | 1.219 | 0.352 | 0.350 | 0.458 | -0.762 (-0.929 to -0.659) | 0.965 | 0.354 | 0.345 | 0.385 | -0.58 (-0.721 to -0.456) |

S<sub>k</sub>: contribution of each characteristic to GINI coefficient
G<sub>k</sub>: GINI coefficient for each characteristic
R<sub>k</sub>: Correlation between GINI coefficient with values of each sociodemographic characteristic
Share: Absolute contribution to inequality= (S<sub>k</sub>\*G<sub>k</sub>\*R<sub>k</sub>/ GINI Value)

In the Gini index decomposition analysis at six months after the start of fourth dose vaccination campaign, the contribution of each sociodemographic characteristic to total inequality (Sk) was higher in white or mestizo ethnic group (Sk= 3.03), people living in rural (Sk= 3.11) or urban areas (Sk= 2.76), as well as the age group between 30 and 64 years (Sk= 3.20), and for men and women (Sk= 2.92 and Sk= 2.94, respectively). In addition, it was identified that the GINI coefficient for each sociodemographic characteristic (Gk) was higher for the Aymara (Gk= 0.92), Afro-Peruvian (Gk= 0.61), and Quechua (Gk= 0.53) ethnic groups, as opposed to the white or mestizo group (Gk= 0.34). Likewise, the correlation between the GINI index and the values of sociodemographic characteristics (Rk) showed high positive values for inhabitants with university or postgraduate education (Rk= 0.44), for people living in urban areas (Rk= 0.28) and for women in provinces (Rk= 0.17). Moreover, high negative values were identified for those living in rural areas (Rk= -0.28), population with elementary education (Rk= -0.36), and those with no education or only preschool education (Rk= -0.32). These trends remained consistent in the GINI index decomposition analysis at twelve months after the start of fourth dose vaccination campaign, however the values were low. Furthermore, another important factor in the inequality in vaccination coverage was the percentage of positivity in provinces; both six (Sk= 1.22, Gk= 0.35, and Rk= 0.35) and twelve (Sk= 0.97, Gk= 0.35, and Rk= 0.35) months after the beginning of the third dose vaccination campaign (**Table 2**).

Regarding the fourth dose immunization campaign at six months, the GINI coefficient of the Lorenz curve was 0.33 (95%CI: 0.30 to 0.35), indicating that the provinces with a higher percentage of white or mestizo inhabitants had higher access to vaccination (**Figure 2C**). Twelve months after the start of the campaign, it remained at 0.31 (95%CI: 0.28 to 0.33), then vaccination coverage inequalities persisted (**Figure 2D**).

**Figure 2.**
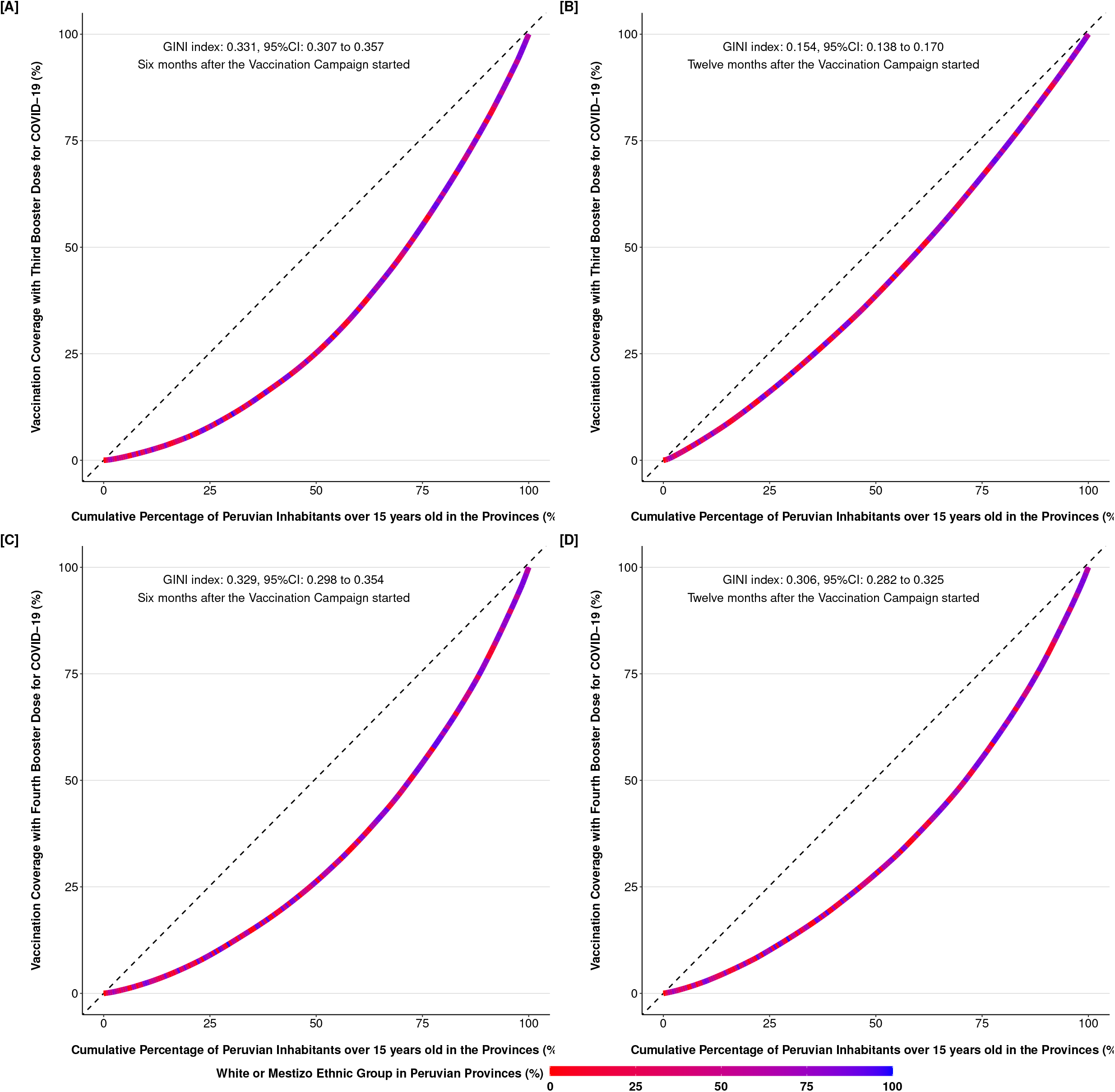
Lorenz curves to evaluate the inequality in coverage with booster doses for COVID–19 in the provinces of Peru at six and twelve months from the beginning of the vaccination campaign

In Peru, the Quechua and Aymara ethnic groups are concentrated in its southern regions. Besides, the inhabitants who identified themselves as white or mestizo are focused on the northern regions. Coincidentally, in the latter provinces, there was a higher percentage of people vaccinated with COVID-19 at six and twelve months after the start of the third dose immunization campaign. Additionally, the distribution of positivity in provinces did not change significatively at six and twelve months after the beginning of third (13.94 vs 17.33, respectively) and fourth (18.78 vs 19.33, respectively) dose immunization campaign (**Figure 3**).

**Figure 3.**
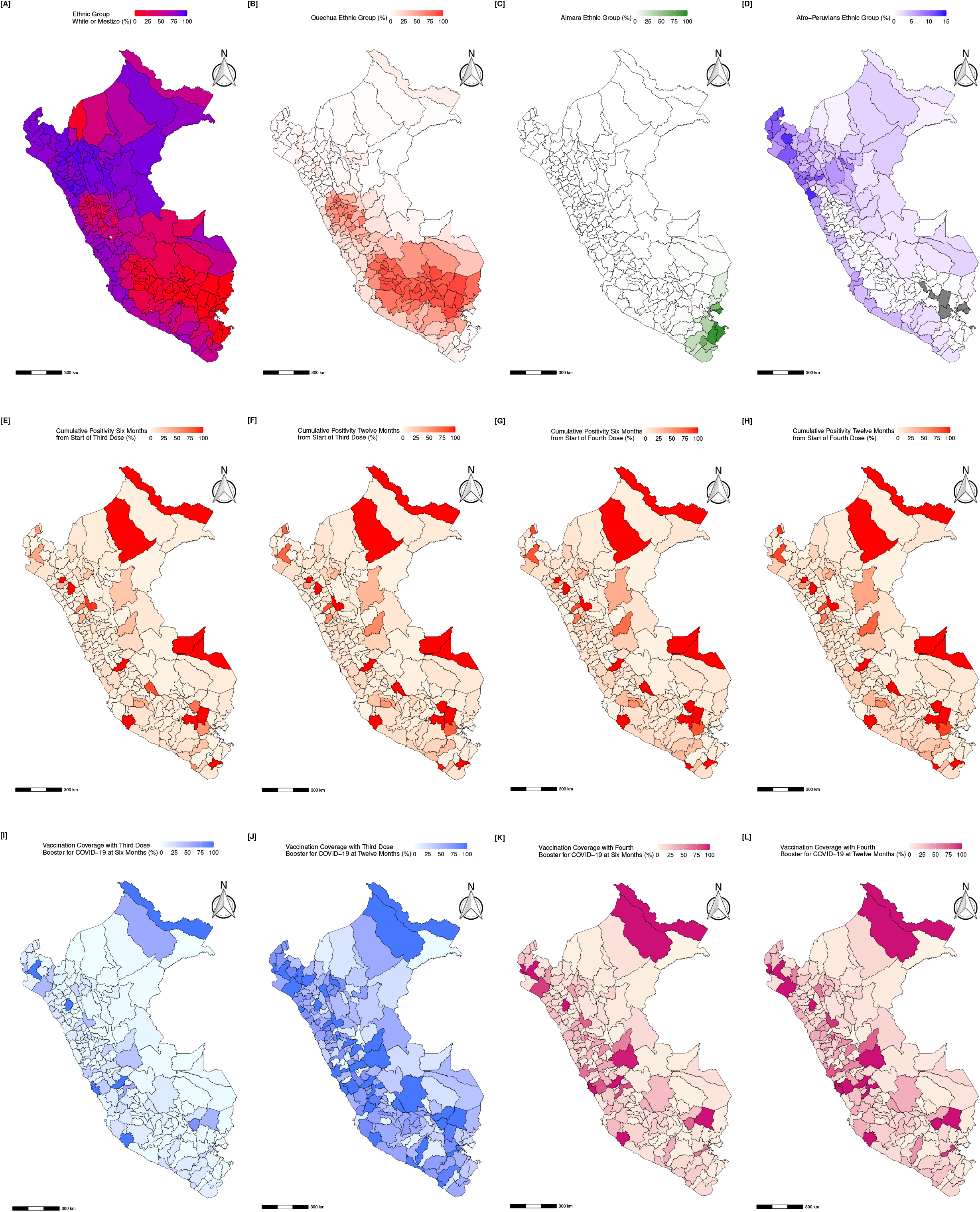
Geographical distribution of ethnic groups, cumulative COVID–19 positivity rate, and vaccination coverage with booster doses for COVID–19 at six and twelve months after the start of each immunization campaign

## DISCUSSION

This study examined vaccination coverage disparities using COVID-19 booster doses in 196 Peruvian provinces. It was discovered that increasing vaccination coverage in the provinces reduced the value of the GINI index more in the third dose than in the fourth dose of the immunization campaign. This finding could imply that immunization campaign strategies are less effective in reducing disparities in vaccination coverage (21,22), which could be attributed to a greater influence of sociodemographic gaps, a perception of a lower risk of infection or death from COVID-19, or an ineffective campaign communication strategy (22,23,24). It currently prioritizes older age groups, so the inequality attributed to this sociodemographic condition is understandable (25).

In the decomposition analysis of the GINI index to evaluate inequality in vaccination coverage by sociodemographic conditions. The most important sources of inequality in vaccination coverage were ethnicity, place of residence, educational level, age group, and gender. Provinces with a higher number of non-white or mestizo ethnic groupings, such as Aymara, Afro-Peruvian, or Quechua populations, had poorer vaccination coverage and higher levels of inequality than provinces with a higher proportion of white or mestizo inhabitants. These ethnic groups often face additional sociodemographic challenges, such as language barriers and discrimination, contributing to distrust and hesitancy towards vaccination (8,26,27). Moreover, factors like a lack of incentives and the belief in natural immunity resulting from previous COVID-19 infections may also contribute to vaccine hesitancy within these communities (28).

The study also found that education is a key inequity driver in vaccination coverage. In provinces with higher levels of education, there was a higher adherence to the COVID-19 vaccine due to better awareness of the risks of the disease, spotting misinformation, and making informed vaccination decisions (29,30). They also had better access to medical care resources and services. But it has been found that education might decrease vaccine coverage in higher-income countries (31). Higher-educated people are more vaccine-hesitant. Due to their greater access to knowledge, these individuals may be more skeptical of booster vaccination against COVID-19 (32).

In addition to the aforementioned sociodemographic factors, the study identified geographic location as a significant contributor to COVID-19 booster vaccine coverage inequalities. Provinces with higher rural populations faced significant challenges in guaranteeing equitable access to healthcare resources, resulting in increased inequality (33). Rural locations provide problems in properly giving booster immunizations, such as limited healthcare infrastructure and specialists, insufficient transportation networks, and significant distances between villages, all of which reduce immunization coverage (33,34). This inequality in access reinforces inequality in rural and urban areas, where people are more likely to be vaccinated.

Although the effectiveness of booster doses has been debated because of the lower risk of COVID-19 infection or mortality and the lack of consensus across countries on the ideal number or types of booster doses, in the administration of a third booster dose, according to vaccination coverage increases the increase of COVID-19 positive rate was low compared to previous months (35). On the other hand, the fourth booster dose immunization campaign resulted in a more significant increase in the COVID-19 positive rate. This finding shows the real risk of low coverage with booster doses because, in these regions, outbreaks of COVID-19 can originate, allowing the development of variants with the possibility of evading the immune response generated with the current vaccines (36).

The study emphasizes the vaccination programs on ethnicity groups, area of residence, and educational level to address sociodemographic inequalities related to booster vaccination coverage. However, this study has an ecological approach and was developed with data from the 2017 census and national open data platforms, which can limit the generalization of findings at the individual level. Additionally, the study evaluated vaccination coverage in inhabitants aged 12 years or older, which may affect the accuracy of the estimates due to variations in the census data.

In conclusion, the study highlights the importance of addressing sociodemographic inequalities in booster dose vaccination coverage for COVID-19, particularly with geographic location, ethnicity, and educational level. The research shows that provinces with higher proportions of Aymara, Afro-Peruvian, Quechua, or other ethnic groups face greater inequality in vaccination coverage than regions with higher proportions of white or mestizo inhabitants. Addressing these sociodemographic conditions is essential for improving vaccination campaigns, especially given the stagnant increase in vaccination coverage for the fourth dose.

## Data Availability

The data used in the study are available in the National Institute of Statistics and Informatics platform (https://censo2017.inei.gob.pe/resultados-definitivos-de-los-censos-nacionales-2017/), Open Data National Platform in Peru (https://www.datosabiertos.gob.pe/dataset/casos-positivos-por-covid-19-ministerio-de-salud-minsa), and website of the Ministry of Health (https://www.minsa.gob.pe/reunis/data/vacunas-covid19.asp).

https://www.datosabiertos.gob.pe/dataset/casos-positivos-por-covid-19-ministerio-de-salud-minsa

https://www.minsa.gob.pe/reunis/data/vacunas-covid19.asp

## Author contributions

CIE and DRB participated in the conception and design of the study; CIE, and GTS participated in data collection, while CIE participated in data management and analysis. All authors participated in the interpretation of the results, the writing and review of manuscript. Also, all authors approved the final version of the manuscript.

## Acknowledgements

None.

